# The majority of the variation in COVID-19 rates between nations is explained by median age, obesity rate, and island status

**DOI:** 10.1101/2021.06.14.21258886

**Authors:** Joseph B. Fraiman, Ethan Ludwin-Peery, Sarah Ludwin-Peery

**Author notes:** corresponding author at < >. These authors contributed equally to this work (Joseph B. Fraiman, Ethan Ludwin-Peery). < >.

## Abstract

Since the World Health Organization declared SARS-CoV-2 to be a global pandemic on March 11, 2020, nearly every nation on earth has reported infections. Incidence and prevalence of COVID-19 case rates have demonstrated extreme geospatial and temporal variability across the globe. The outbreaks in some countries are extreme and devastating, while other countries face outbreaks that are relatively minor. The causes of these differences between nations remain poorly understood, and identifying the factors that underlie this variation is critical to understand the dynamics of this disease in order to better respond to this and future pandemics.

Here, we examine four factors that we anticipated would explain much of the variation in COVID-19 rates between nations: median age, obesity rate, island status, and strength of border closure measures. Clinical evidence suggests that age and obesity increase both the likelihood of infection and transmission in individual patients, which make them plausible demographic factors. The third factor, whether or not each country is an island nation, was selected because the geographical isolation of islands is expected to influence COVID-19 transmission. The fourth factor of border closure was selected because of its anticipated interaction with island nation status.

Together, these four variables are able to explain a majority of the international variance in COVID-19 case rates. Using a dataset of 190 countries, simple modeling based on these four factors and their interactions explains more than 70% of the total variance between countries. With additional covariates, more complex modeling and higher-order interactions explains more than 80% of the variance. These novel findings offer a solution to explain the unusual global variation of COVID-19 that has remained largely elusive throughout the pandemic.

## 1. Introduction

On March 11, 2020 the Director General of the World Health Organization (WHO) declared the novel Coronavirus (COVID-19) as a pandemic (WHO, 2020). Since then, COVID-19 has spread to nearly^1^ every nation in the world, but some nations have been more severely impacted than others, with thousand-fold differences between nations in their infection rates per capita. At the start of the pandemic, most predictions held that wealthier, western nations would do the best at controlling the spread of the virus and mitigating its impacts (Brauer et al, 2020; Nuwagira & Muzoora, 2020; Hodkinson et al., 2020; Myre, 2020; Ali & Ali, 2020) because of their wealth and access to high-quality healthcare, but in fact we have seen the opposite, with developed nations generally suffering large numbers of cases per capita, and poorer nations generally experiencing much more limited outbreaks.

There has been great interest in understanding the causes of the large disparity in COVID-19 cases between nations to explain today’s worldwide patterns, with many lay press articles exploring this (Beech et al., 2020; Mukherjee, 2021; Leonhardt, 2021), following up on the large numbers of exploratory studies examining this (Van Damme et al., 2020; Skorka et al., 2020; Chakraborti et al., 2021). Multiple hypotheses have been proposed to explain the causes of this great variability in infection rate, including population density, age, timing of lockdown, temperature, urban density, containment policies, obesity rates, GDP, pollution, non-pharmaceutical interventions (travel restriction, limited event sizes, mask mandates, etc.), smoking rates, humidity, latitude, and collectivism (Singh et al., 2021, Chaudhry et al., 2020, Yu et al., 2021, Iyanda et al., 2020, Wu et al., 2020, Duhon et al., 2021, Hradsky & Komarek, 2021, Leung et al., 2020, Nguimkeu & Tadadjeu, 2021). Despite this, the major causes of the worldwide pattern remain elusive.

Some proposed factors are more closely related to the demographics of the population, while others have more to do with national attributes and policy. We suspect that factors in both categories play a role. Below we propose two demographic factors, one geographic factor, and one policy decision that may explain much of the variation.

### 1.1. Demographics

#### 1.1.1. Superspreaders

COVID-19 transmission is largely driven by superspreaders, with 20% of those infected responsible for 80% of transmission (Miller et al., 2020; Wu et al., 2020; Endo, 2020). Genome mapping studies of SARS-CoV-2 variants indicate that superspreaders are the main driver of COVID-19 cases (Gomez-Carballa et al., 2020), and modeling suggests that reducing superspreader events before widespread infection would cause outbreaks to fizzle out (Althouse et al., 2020). Given the critical role superspreaders play in wide-spread infection, nations with fewer superspreaders would have lower COVID-19 transmission and therefore lower rates of infection. As a result, anything that reduces the number of superspreaders or the prevalence of superspreader events will appear to have an outsized impact on the size and severity of outbreaks.

An individual’s likelihood to transmit COVID-19 depends on two factors. The first factor is the likelihood that they will become infected, as one must be infected in order to further transmit disease. Increased likelihood of infection could occur from increased contact with those with COVID-19, or increased susceptibility to infection following contact. The second factor is the likelihood that, if infected, they will transmit to others. Increased likelihood of transmission would be expected in those who have more contact with others, or increased likelihood of transmission with each contact (Kupferschmidt 2020).

If the likelihood of either factor is low — that is, if an individual is unlikely to become infected, or in becoming infected, is unlikely to transmit the infection to others — an individual is unlikely to spread the infection at all. If one of these factors becomes likely, then the person has an increased chance to spread the disease. And if both factors are likely — if someone is likely to catch the disease, and having caught it, likely to transmit it — then that person is more likely to become a superspreader.

If a nation does not have a large number of individuals who can become superspreaders, COVID-19 transmission is unlikely to grow exponentially. (Endo, 2020) As a result, when trying to explain the differences in spread between countries, we should pay close attention to aspects of policy and demographics that are likely to influence either of these factors, and special attention to anything that might influence both. Any factor that makes people both likely to catch COVID-19, and likely to spread it, will have a disproportionate impact on the variance in case rates worldwide.

Because of these dynamics, it is important to identify the factors that are likely to increase the likelihood of infection or transmission, and particularly the factors that increase both infection and transmission.

#### 1.1.2. Age

Of the previously proposed explanations for the variance between countries, differences in median age between nations is among the most promising. Nations with younger populations generally have lower rates of COVID-19, while the rates for nations with older populations are generally higher (Dowd et al., 2020). This is to be expected, as older adults are more susceptible to COVID-19 infection and more likely infectious if infected (Madewell et al., 2020; Jing et al., 2020).

Increased susceptibility with increasing age has been most clearly demonstrated using secondary attack rate (SAR) studies, in which most studies have found when exposed to a household contact who tested positive for COVID-19, children have decreased susceptibility compared with adults (Madewell et al., 2020). Multiple other studies have shown the risk of symptomatic infection in adults increases with age, with individuals over 50 generally showing a 2- to 3-fold increase in susceptibility to infection following a COVID-19 contact (Leung et al., 2020).

For an individual infected with COVID-19, infectivity increases with age. A review of household studies found adults were approximately five times more likely to be the primary case than children (54% vs 9%, respectively) (Zhu et al., 2020). The difference in infectivity continues to increase with age. For example, a household contact SAR study found individuals under 20 had a SAR of only 5.3%, while individuals 20-59 had a SAR of 14.8% and individuals over 60 had a SAR of 18.4% (Jing et al., 2020). Unsurprisingly, children are unlikely to be COVID-19 superspreaders (Munro et al., 2020).

While the exact cause of why infectivity increases with age is not fully understood, it is likely partially dependent on how age increases the severity of symptoms (Matsushita et al., 2020), which increases the likelihood of transmission (Luo et al., 2020). Secondary attack rates (SAR) from individual asymptomatic infections are only 0.3%, while cases with mild symptoms have a SAR of 3.3% (a 10-fold increase), and cases with severe symptoms have a SAR of 6.2% (a 20-fold increase compared to asymptomatic infections).

Because age increases the likelihood of catching COVID-19 if exposed, and transmitting it if infected, it seems likely that median age is an important factor in explaining the variance between nations, and previous research has supported this hypothesis.

#### 1.1.3. Obesity

Like age, obesity is a well-established risk factor for increased severity of disease in general (Gao et al., 2020; Kalligeros et al., 2020). In addition to increased severity, obesity increases individuals’ susceptibility to COVID-19. In those who have been tested for COVID-19, obesity has been found to increase the likelihood of a positive result by approximately 75% (de Lusignan et al., 2020). An analysis of over 400,000 individuals found obesity increased the relative risk of 2.32 for testing positive for COVID-19 infection (Ho et al., 2020). While individuals of normal weight (BMI < 25) in this study were 34% of the population, they were only 21% of the total number of infections.

Whether obesity increases the likelihood of transmission in those who are infected has not been thoroughly investigated. However, obesity clearly increases the severity of COVID-19 symptoms (Gao et al., 2020; Kalligeros et al., 2020). Increased symptom severity increases COVID-19 infectivity, and this likely contributes to the increased infectivity observed with age (Luo et al., 2020). It is also likely that increased severity of symptoms in obese individuals increases infectivity, which would increase transmission.

#### 1.1.4. Demographic Interactions

In addition, there is evidence that the effects of age and obesity in combination are more than the sum of their parts. A recent paper examining exhaled aerosol particles found that approximately 20% of individuals were the source of 80% of total particle production (Edwards et al., 2021). In the group producing 80% of aerosol particles, the study found a positive correlation between particle production and both age and BMI. In addition, when the team multiplied age and BMI to create “BMI-years”, they found that the individuals with the lowest BMI-years exhaled significantly less bioaerosol than the individuals with the highest BMI-years.

However, demographic factors are not the only factor that should be expected to impact COVID-19 rates. National policy and geographic features may play a role as well.

### 1.2. National Features

#### 1.2.1. Islands

There are multiple reasons to expect that island nations would be able to use their geographical isolation to protect against COVID-19. It is said that many national borders are little more than lines in the sand. Sometimes, they follow the course of a river or the peaks of a mountain range. The borders of islands are categorically different. Islands are physically isolated from the rest of the world. This may be part of why islands tend to have relatively high COVID-19 testing effectiveness; Kuster & Overgaard (2021) defined a COVID-19 Testing Index (a measure of a nation’s approach to testing) derived from epidemiological indicators of testing and found that island nations generally had a high testing index, with islands accounting for more than half of the top ten countries in terms of testing effectiveness. Multiple studies have suggested that island nations have a geographical advantage protecting them from COVID-19 entering the country (Baum, et al., 2021, Mayer, & Lewis 2020), and machine learning approaches have previously identified island status as a likely predictor of COVID-19 rates (Li et al., 2021).

Even prior to COVID-19, island nations were considered an ideal refuge to escape deadly pandemics (Boyd et al., 2017, Turchin & Green 2019), and they have an impressive track record. In the influenza pandemic of 1918, one of the deadliest pandemics in history, the only regions that were entirely spared were American Samoa (Killingray & Phillips, 2003), St. Helena, Tristan da Cunha, and parts of Iceland, all island nations that escaped the pandemic through “rigorous maritime quarantines” (Vollmer & Wójcik, 2017).

Even among the islands that were eventually infected, many managed to hold the virus at bay for months by imposing similar quarantines (McLeod et al., 2008, Markel et al., 2006). Islands such as Fiji, Madagascar, Tasmania, Nauru, Tonga, Guam, Western Samoa, and New Zealand kept themselves out of the first wave of the pandemic entirely, though they were eventually infected late in the second wave. A “vigorous” maritime quarantine protected Australia for almost a year, until January 1919. Notably, while maritime quarantines did temporarily or even totally protect a small number of island nations, “quarantines proved useless almost everywhere else” (Patterson & Pyle, 1991).

#### 1.2.2. Border Closure Policy

Islands’ unambiguous border strength likely allows border closure policies to have a greater impact for islands than for mainland nations. Global analyses tend to find that border closures have only limited effectiveness (Mallapaty, 2020, Grepin et al., 2021, Chinazzi et al., 2020), but studies of individual island nations have found border closures to be highly effective in countries such as Australia (Adekunle et al., 2020), Hong Kong (Cowling et al., 2020), and the Pacific Islands (Leal Filho et al., 2020). In Caribbean island nations, strict border closures were associated with reduced COVID-19 rates, and when nations reduced their levels of closure, there was a temporal association with an increase in COVID-19 cases (Murphy et al 2020).

The only large cross-country study examining the effectiveness of various non-pharmaceutical interventions (NPIs) on COVID-19 growth rate found that international movement restrictions were the only NPI to approach significance (*p* = 0.062) (Duhon et al., 2020). This paper did not examine the interaction of island status and travel restrictions in their analysis. If such a relationship exists, they would have missed it, and this would explain why they found only limited evidence for the efficacy of border control measures. Possibly the best evidence that exists that the unique geographical isolation of island nations along with strong border closure can be prevent COVID-19 infections is that the only five nations in the world that are universally accepted to have had avoided any COVID-19 infections are Palau, Tuvalu, Kiribati, Nauru, and Tonga, all sovereign island nations who have maintained strong border closures throughout the pandemic.

Given the geographical uniqueness of island nations, we expected that island status would be a major factor in explaining the differences in COVID-19 rates between nations. Because this factor is likely to change the dynamics of COVID-19 transmission, we further expect that there will be statistical interactions between this and other factors.

### 1.3. Hypotheses

Based on the evidence outlined above, we expect that median age, obesity rate, island status, and the degree of border control measures will be major factors in explaining the differences in COVID-19 rates between nations. To test this hypothesis, we performed a series of regression analyses based on data from March 2021, one year after the WHO declared the pandemic in March of 2020. The influences of these variables may manifest as main effects, interaction effects, or both. We specifically anticipate an interaction in the case of island status and border closure, as we think there is reason to believe that border closure measures are more effective for islands than for mainland nations.

#### 1.3.1. Previous studies

Of the four critical variables, only age has consistently been identified to be associated with COVID-19 case rates (Iyanda et al., 2020,Nguimkeu, & Tadadjeu, 2020, Gangemi, Billeci & Tonacci, 2020). For the other three critical variables, the evidence is more mixed or inconsistent. Some studies have found obesity rate to be associated with COVID-19 rates (Leung et al., 2020, Yu et al., 2020, Chaudry et al., 2020), but other studies have not observed the association (Li et al., 2021). Border closure has been included in some models, but generally has not been found to be a significant predictor of COVID-19 case rates in cross-country analyses (Duhon et al., 2021). One data-driven analysis of 77 potential predictors previously identified island status as one likely predictor of COVID-19 rates (Li et al., 2021). However, the authors of this paper interpreted this as an effect of climate (“the weather is humid in island countries”) rather than the effect of the unique border quality of islands, and did not investigate any statistical interactions.

The reason the variables of obesity, island status, and border closure may not have been consistently identified as being associated with COVID-19 rates, is a lack of studies examining the interactions between these four variables. The hypothesis predicts that age and obesity strongly influence COVID-19 rates in mainland nations, and in islands without strong border closure, while the full impact of border closure on COVID-19 rates will only be fully appreciated in island nations, and likely have little impact in non-island nations.

In addition to our critical variables, other variables that have been previously identified to be consistently associated with COVID-19 rates were included as covariates.

Our first main covariate is GDP per capita at purchasing power parity, or GDP per capita PPP. This was included because it is a general measure of wealth and standard of living, and we want to correct for this as some of our critical variables are associated with the same. In addition, measures of GDP have previously been found to be positively associated with COVID-19 case rates (Gangemi, Billeci & Tonacci, 2020, Yu et al., 2020).

Nations with more interpersonal contact should have greater COVID-19 transmission, leading to increased COVID-19 cases per capita. Surrogates for level of interpersonal contact include higher population per square mile (population density), and percent of population living within cities (urban population), both of which have been found to be related to COVID-19 cases per capita (Singh et al., 2021, Yu et al., 2020, Nguimkeu & Tadadjeu, 2021). These variables have also been associated with higher COVID-19 growth rate (Duhon et al., 2021, Leung et al., 2020, and total deaths (Hradsky & Komarek, 2021). For these reasons, we also included population density and urban population as covariates in our analyses.

#### 1.3.2. Exponential Dynamics

Pandemics have exponential dynamics. As reproduction number (R0) in a population rises above 1, the infection not only spreads, but spreads exponentially.

Factors that predict case numbers are multiplicative rather than additive. If there are two factors that influence the spread of the pandemic, the outcome for a country with both factors will be more than twice as bad as for a country that has only one factor. If there are three factors, a country with any two factors will do much worse than a country with only one of the factors, and a country with all three will do even worse, and so on, in a superadditive manner.

This leads us to two basic predictions for our modeling. The first is rather straightforward, and widely recognized in similar analyses (e.g. Duhon et al., 2021, Hradsky & Komarek 2021), which is that we should expect our dependent variable, COVID-19 cases per million, to follow a lognormal distribution, because the differences in case rates between countries should be governed by exponential dynamics.

The second is alluded to above, that we should also expect multiplicative dynamics in our predictors. In a multiple regression, this will be expressed as significant interaction terms. As a result, after examining main effects, we will examine two- and three-way interactions between our variables of interest.

## 2. Methodology

### 2.1. Data Collection

All data extracted was publicly available and obtained via <u>ourworldindata.com</u>, other than island nation status, which was obtained from the nations online program (nations online program, 2021), and GDP per capita PPP, which was obtained from the CIA World Factbook. median age was missing for two nations and was completed using data from the World Bank. Minimum border closure policy, henceforth border closure, was calculated as the minimum level of border closure measures between April 1st 2020 and April 27, 2021, the date at which the analysis was performed. The border closure variable could be any discrete value between 0 and 4, with 4 being the most strict border closure regulation and 0 being no regulation (see Box 1 for details of border closure ranking definitions). Border closure ratings were missing for nineteen nations; we filled this by looking up the official government statements and the date(s) on which those statements were made for each of the missing nations, and the first author coded these by hand. All other data was collected in March 2021, a year after COVID-19 was declared a pandemic. More detail on our sources can be found on <u>the OSF page for this project</u>.

#### Box 1

Border Closure Measure per <u>Blavatnik School of Government, University of Oxford</u>

0 - No measures

1 - Screening

2 - Quarantine arrivals from high-risk regions

3 - Ban on high-risk regions

4 - Total border closure

Table 1 presents basic descriptive statistics of our variables of interests for a total of 190 countries. The variables island status and border closure are not included as they are categorical variables.

**Table 1.** Summary Statistics.

| Variable | Observations | Mean | Std. Dev. | Min | Max |
| --- | --- | --- | --- | --- | --- |
| **COVID-19 Cases per mil.** | 190 | 24,787.17 | 31,017.42 | 0.00 | 153,368.28 |
| **GDP per capita PPP** | 189 | 19,935.79 | 20,498.79 | 752.00 | 114,482.00 |
| **Population Density** | 188 | 201.66 | 618.61 | 2.00 | 7,916.00 |
| **Urban Population (%)** | 189 | 0.58 | 0.23 | 0.13 | 1.00 |
| **Median Age** | 190 | 28.93 | 8.82 | 14.90 | 46.30 |
| **Obesity Rate (%)** | 190 | 0.20 | 0.11 | 0.02 | 0.61 |

We were concerned about whether differences in the accuracy of reporting COVID-19 cases might potentially influence our results. To examine the potential influence of under-reporting, we applied a model (Favero, 2020) which uses a nation’s positivity rate to create an adjusted COVID-19 case rate. The adjusted case rate increases the confirmed case rate across all nations by an extra 0.1 to 0.2 cases for every percentage increase in positivity rate to help account for underreporting. We found that the adjusted and unadjusted COVID-19 case variables were highly correlated, *r*(100) = .995, *p* < .001, and were even more highly correlated when both were log-transformed, *r*(100) = .999, *p* < .001. Because the two were so highly correlated, the results of all analyses will be nearly identical regardless of which measure is used. Because it was not possible to adjust case rates for all countries, using adjusted cases would reduce our sample size from 190 to only 102 countries, eliminating much of the variance we want to try to explain. As a result we elected to use unadjusted COVID-19 cases throughout.

### 2.2. Regression Analysis

As expected, the distribution of COVID-19 cases per million was heavily skewed, approximating a lognormal distribution. To account for this, we log-transformed this variable and used the resulting log COVID-19 cases per million (henceforth, log cases) as the dependent variable in all analyses.

Before conducting our regression modeling, we examined the correlations between all variables of interest (minus the variable island, which is categorical). All correlations appear in Table 2.

**Table 2.** Zero-order Pearson correlations.

|  | **Log COVID Cases per M.** | **GDP per capita PPP** | **Population Density** | **Urban Population (%)** | **Median Age** | **Obesity Rate (%)** |
| --- | --- | --- | --- | --- | --- | --- |
| **Log COVID Cases per M.** | – | 0.465 *** | 0.040 | 0.475 *** | 0.512 *** | -0.006 |
| **GDP per capita PPP** |  | – | 0.289 *** | 0.622 *** | 0.720 *** | 0.265 *** |
| **Population Density** |  |  | – | 0.141 | 0.121 | -0.075 |
| **Urban Population (%)** |  |  |  | – | 0.556 *** | 0.486 *** |
| **Median Age** |  |  |  |  | – | 0.330 *** |
| **Obesity Rate (%)** |  |  |  |  |  | – |
| Note: * p < .05, ** p < .01, *** p < .001 |  |  |  |  |  |  |

The majority of our variables of interest were weakly to moderately correlated, and most of these relationships are statistically significant (Table 2). To ensure that our variables provide unique or independent information in the regression model, we calculated the variance inflation factor (VIF) of all covariates using the vif() function from the car (Companion to Applied Regression) package in R (Fox & Weisberg 2019). There is no formal cutoff for interpreting VIF, and various cutoffs are used for determining what magnitude of VIF indicates concerning levels of multicollinearity. Cutoffs can be as high as 10, but similar work has tended to use more conservative cutoffs for VIF, using for example a cutoff of 5 (Duhon et al., 2021) or 3 (Iyanda et al., 2020). The VIF of all covariates was less than this most conservative cutoff, with the largest VIF being only 2.67. As a result there is evidence of an extremely minor degree of multicollinearity, but not enough to be of any concern.

The main analyses were conducted using multiple regression. Analyses consisted of a multiple linear regression model of the form:

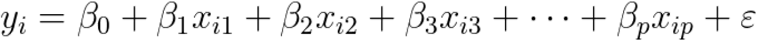

where, for *i* = 1 to *n* observations:

*y*_*i*_ = dependent variable

*β*_0_ = constant (y-intercept)

*β*_j_(j from 1 to p) = coefficient or slope for each covariate

*x*_*ij*_ = covariates, where *j* ranges from 1 to p, p being the total number of covariates

*ε* = the error term (residuals)

Before fitting models, all variables were centered and standardized.

The raw data and the analysis scripts can be found on <u>the OSF page for this project</u>.

#### 2.2.1. Covariates

To begin with, we fit a model that only included our covariates and didn’t include any of our critical variables. This model appears in Table 3 as Model 1. Together, these variables were able to explain 27.0% of the variance. GDP per capita PPP and Urban Population were statistically significant predictors in this model; Population Density is not.

**Table 3.** Model Coefficients.

|  | Intercept | GDP per<br>capita PPP | Urban<br>Population<br>(%) | Population<br>Density | Island | Border<br>Closure | Median<br>Age | Obesity<br>Rate (%) | Island x<br>Border<br>Closure | Median<br>Age x<br>Obesity<br>Rate | Island x<br>Obesity<br>Rate | Border<br>Closure x<br>Obesity<br>Rate | Adjusted R<sup>2</sup> |
| --- | --- | --- | --- | --- | --- | --- | --- | --- | --- | --- | --- | --- | --- |
| **Model 1** | 0.01 | 0.32 *** | 0.29 *** | -0.09 |  |  |  |  |  |  |  |  | 0.270 |
| **Model 2** | 0.20 ** | 0.34 *** | 0.25 ** | -0.01 | -0.76 *** |  |  |  |  |  |  |  | 0.368 |
| **Model 3** | 0.66 *** | 0.37 *** | 0.29 *** | -0.02 | -0.58 *** | -0.29 *** |  |  |  |  |  |  | 0.459 |
| **Model 4** | 0.73 *** | 0.10 | 0.23 *** | 0.02 | -0.59 *** | -0.32 *** | 0.42 *** |  |  |  |  |  | 0.538 |
| **Model 5** | 0.66 *** | 0.09 | 0.29 *** | -0.01 | -0.52 *** | -0.29 *** | 0.43 *** | -0.12 |  |  |  |  | 0.545 |
| **Model 6** | 0.34 *** | 0.02 | 0.11 | 0.04 | 0.12 | -0.06 | 0.30 *** | 0.72 *** | -0.28 ** | 0.14 * | -0.61 *** | -0.15 *** | 0.729 |
| **Model 7** | 0.32 *** |  |  |  | 0.16 | -0.05 | 0.34 *** | 0.79 *** | -0.32 *** | 0.12 * | -0.66 *** | -0.15 *** | 0.716 |

\* p < .05, \*\* p < .01, \*\*\* p < .001

#### 2.2.2. Critical Variables

Having established a baseline fit with the covariates, we went on to see how much additional variance could be explained by our critical variables. All models appear in Table 3.

In Models 2-5, we add the critical variables one at a time. Model 2 adds the Island variable. Island was a significant predictor, β = −0.758, *p* < 0.001, and the addition of this variable explains an additional 9.8% of the variance over the previous model. Island status continued to be an important predictor. Figure 1 shows the differences in log cases between mainland and island nations.

**Figure 1.**
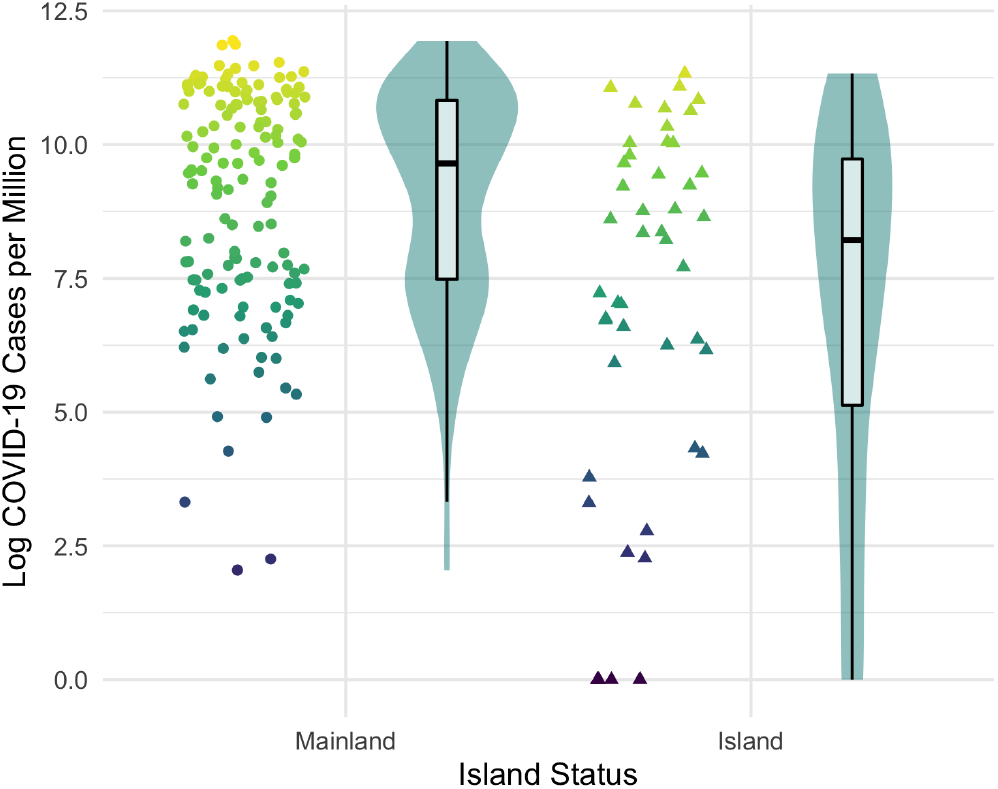
Log COVID-19 Cases per million of Mainland and Island Nations.

Model 3 adds border closure, which is a significant predictor, β = −0.287, *p* < 0.001, and explains an additional 9.1% of the total variance over the previous model. Model 4 adds median age, which is a significant predictor, β = 0.424, *p* < 0.001, and the addition of this variable explains 7.9% of the total variance over the previous model.

Model 5 adds obesity rate, for a model that includes all four critical variables. Obesity rate was not a significant predictor in this model. However, as we see in the next models, obesity rate has multiple significant interactions with the other critical variables. In addition, it’s noteworthy that Model 5, including only the main effects for all variables, explains 54.5% of the variance in log COVID-19 cases.

Anticipating interactions, in the next model we tested two-way interactions involving our critical variables. We started by examining all two-way interactions between the critical variables, and found four that were reliably significant: island x border closure, median age x obesity rate, island x obesity rate, and border closure x obesity rate. These four interactions were included in Model 6. The overall regression equation was significant, F(11, 175) = 46.58, *p* < .001, and this model explained 72.9% of the variance. This is 18.4% more than Model 5, which included only the main effects. The main effects for median age and obesity rate are significant in this model, as are all four interaction terms.

These interaction terms are critically important, and are worth examining further. Figures 2-4 show partial visualizations of Model 6. The figures show unadjusted data, but we note that Model 6 shows that all interactions remain significant when controlling for GDP and other covariates.

**Figure 2.**
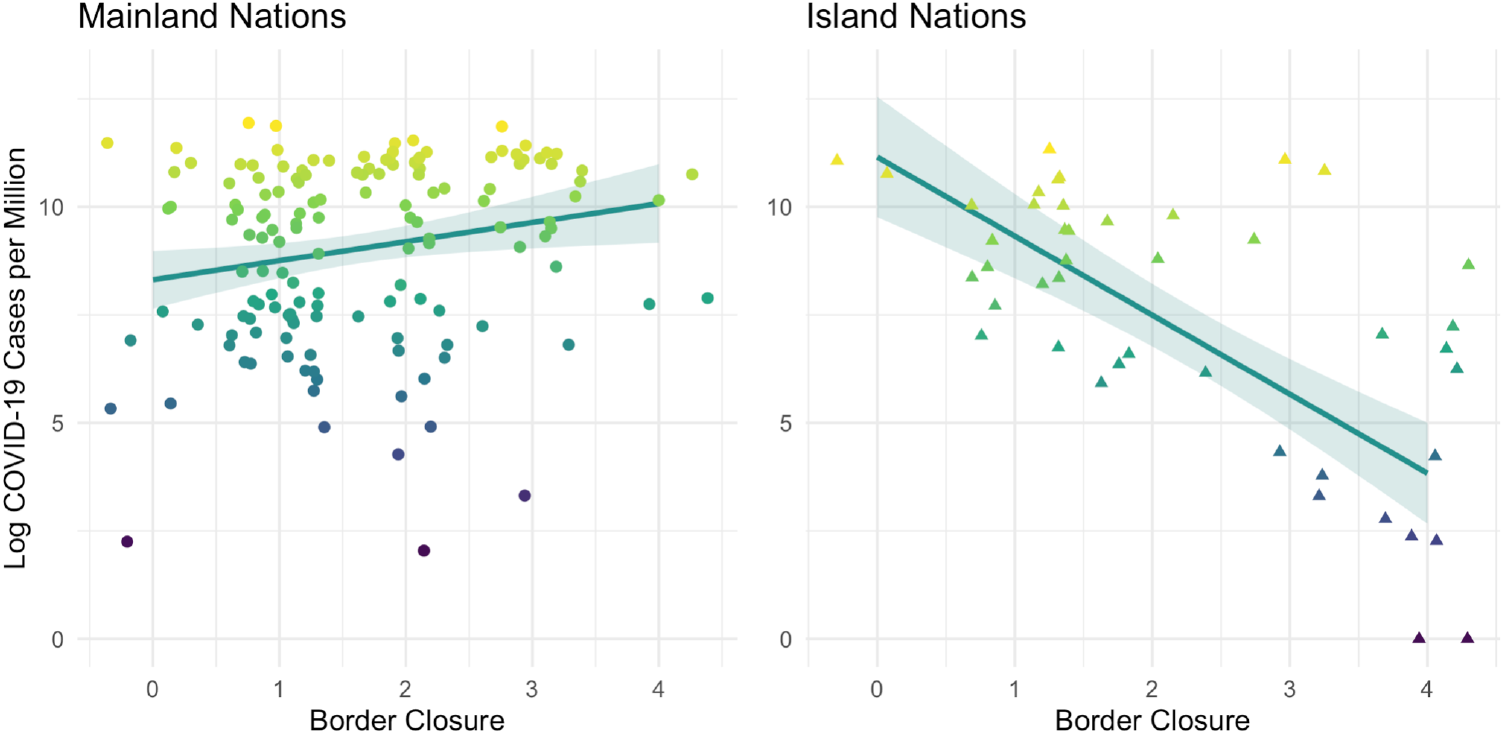
Interaction of Border Closure with Mainland and Island Nations. *Note: Points are jittered to prevent overplotting caused by discreteness in the x variable*.

Figure 2 clearly shows that border closure measures have a different impact on mainland and island nations. Border closure has little to no relation to COVID-19 cases in mainland nations, but for island nations, a greater degree of border closure is associated with lower COVID-19 cases.

Figure 3 shows the strongest interaction term from Model 6, the interaction between obesity rate and island status. In a simple correlation, Obesity doesn’t appear to be associated with COVID-19 case rates (*r* = −0.006, *p* = .933), but plotting this relationship separately for mainland and island nations shows a clear interaction. In mainland nations, there was a distinct positive relationship between obesity and COVID-19 cases, such that nations with higher rates of obesity have higher case rates.

**Figure 3.**
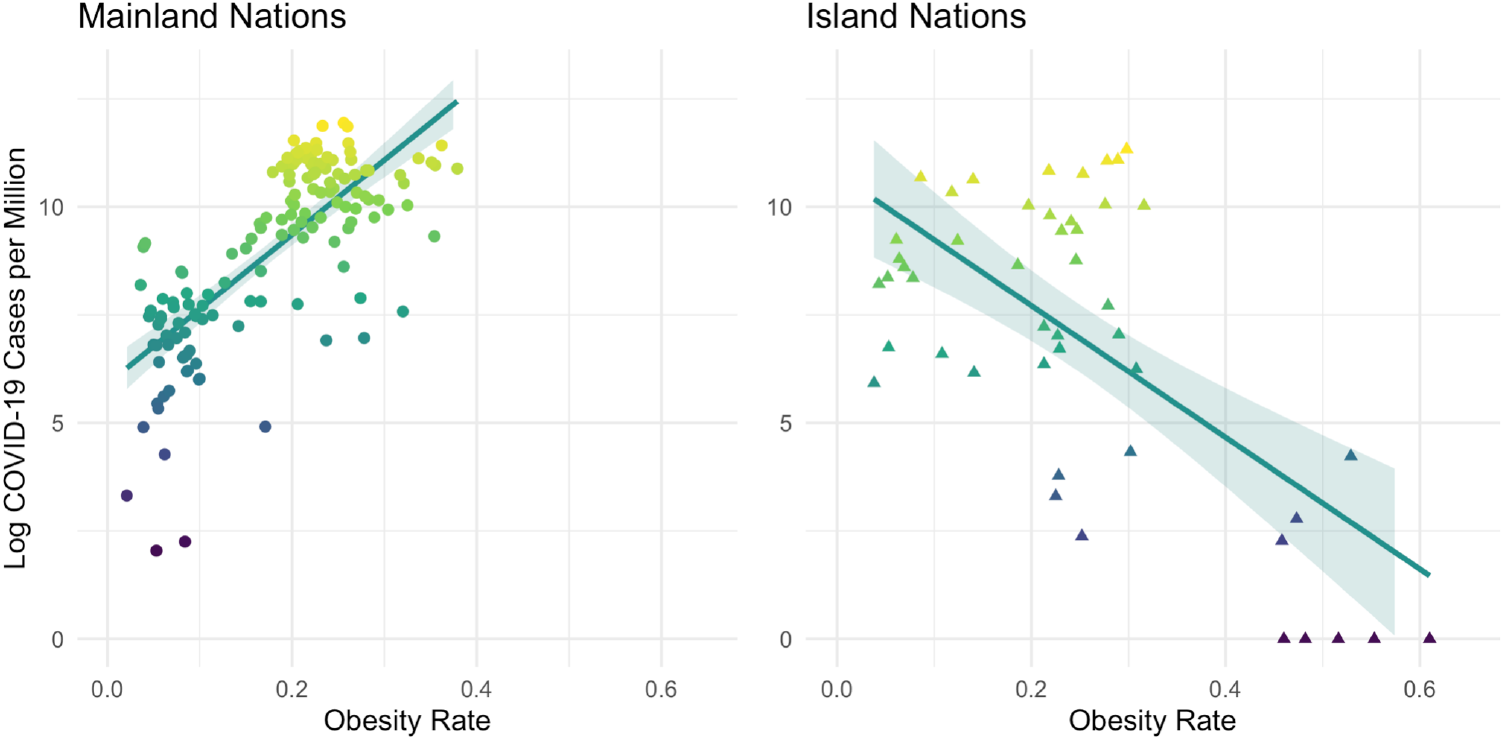
Interaction of Obesity Rate with Mainland and Island Nations.

For island nations, there appears to be a moderate negative relationship, but this may be spurious. This negative relationship appeared to be largely driven by eight island nations with extremely high rates of obesity. In fact, these are the eight most obese nations in this dataset, the only nations with obesity rates above 40%. This is why in the plot of mainland nations there are no points on the righthand third of the figure; there are no mainland nations with an obesity rate higher than 40% in this dataset.

Figure 4 approximates the interaction between Median Age and Obesity Rate. Note the same set of extreme values from Figure 3, and that they are all islands. This emphasizes the importance of controlling for island status and its interactions for understanding the other effects at play in these data.

**Figure 4.**
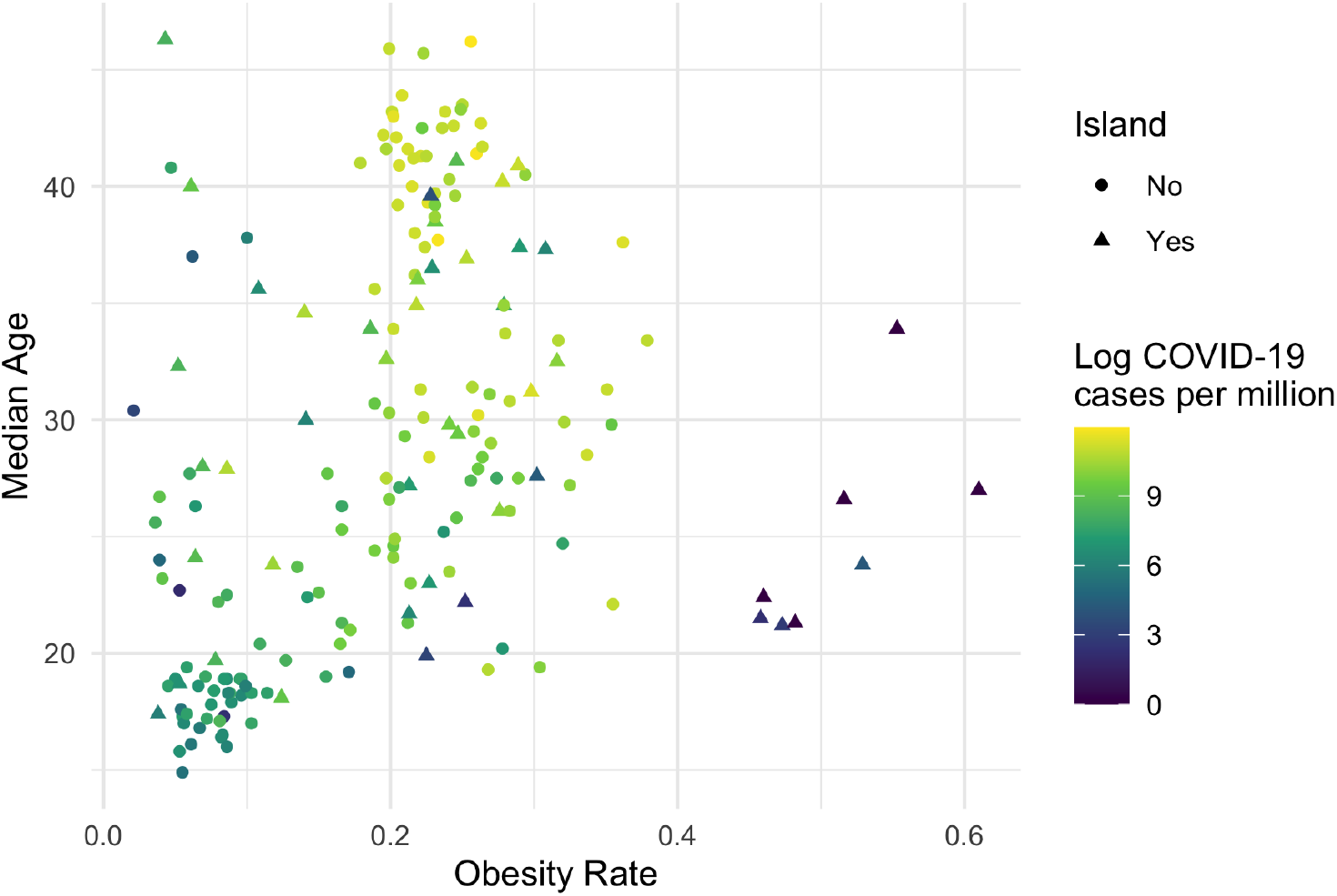
Association of Age and Obesity with Log COVID-19 Cases.

In Model 6, none of the three covariates (GDP per capita PPP, urban population, and population density) are significant. Therefore in Model 7, we fit a model in which we dropped these predictors, and included only our critical variables and their interactions. The regression equation was significant, F(8, 181) = 60.58, *p* < .001. By themselves, these four variables and their interactions were still able to explain a full 71.6% of the variance in log cases between a total of 190 nations. As in Model 6, the main effects for median age and obesity rate are significant, as are all four interaction terms.

#### 2.2.3. Stepwise Regression

As anticipated, we found multiple significant interactions between our critical variables. Because of this success, and because we *a priori* expected there to be many interactions in general, we next examined the full set of two-way and three-way interactions between all variables of interest.

Many of these two- and three-way variables were significant. To help perform model fit with such a large number of predictor variables, we used the stepAIC() function from the MASS package in R (Venables & Ripley, 2002), which selects the best model by AIC, to perform forward and backward stepwise variable selection. Both forms of stepwise variable selection returned similar models with very high model fits: 79.4% for forward selection, and 81.7% for backward selection. Because the backward selection both provided a slightly better fit and included fewer model terms, this is the model we report below.

The overall model was significant, F(43, 143) = 20.34, *p* < .001, and as mentioned above, it explained 81.7% of the variance in log cases between a total of 187 countries.

Forty-four coefficients were retained in the model. The full list is too large to report here, but the twelve significant three-way interactions appear in Table 4. Some of these interactions may be spurious, but many were highly significant (five are *p* < .001) and likely to be robust.

**Table 4.** Significant Three-Way Interactions in Stepwise Model.

| Interaction | Estimate | 95% CI<br>lower | 95% CI<br>upper | Std. Error | t | p |
| --- | --- | --- | --- | --- | --- | --- |
| GDP per capita PPP x Urban Population x Population Density | -1.150 | -1.656 | -0.645 | 0.256 | -4.498 | < .001 *** |
| GDP per capita PPP x Island x Border Closure | 0.355 | 0.138 | 0.571 | 0.110 | 3.234 | 0.002 ** |
| GDP per capita PPP x Island x Obesity Rate | 0.788 | 0.272 | 1.303 | 0.261 | 3.022 | 0.003 ** |
| Urban Population x Population Density x Border Closure | 0.552 | 0.150 | 0.954 | 0.204 | 2.712 | 0.008 ** |
| Urban Population x Population Density x Median Age | 0.814 | 0.170 | 1.457 | 0.326 | 2.500 | 0.014 * |
| Urban Population x Island x Median Age | -0.338 | -0.646 | -0.030 | 0.156 | -2.167 | 0.032 * |
| Urban Population x Median Age x Obesity Rate | -0.259 | -0.407 | -0.112 | 0.075 | -3.475 | < .001 *** |
| Population Density x Border Closure x Median Age | -1.177 | -1.838 | -0.516 | 0.334 | -3.522 | < .001 *** |
| Population Density x Border Closure x Obesity Rate | -0.472 | -0.838 | -0.106 | 0.185 | -2.55 | 0.012 * |
| Population Density x Median Age x Obesity Rate | 0.900 | 0.416 | 1.384 | 0.245 | 3.676 | < .001 *** |
| Island x Border Closure x Obesity Rate | -0.252 | -0.469 | -0.036 | 0.109 | -2.306 | 0.023 * |
| Island x Median Age x Obesity Rate | -0.775 | -1.202 | -0.347 | 0.216 | -3.584 | < .001 *** |

## 3. Discussion

This study identifies four variables — median age, obesity, island status, and border closure — and finds that they are able to explain the majority of variance of COVID-19 rates between nations. Together, a simple model based on these four variables and their two-way interactions explains more than 70% of the variance of COVID-19 rates worldwide. A more complex model using three-way interactions and three additional variables — GDP, population density and urban population — is able to explain over 80% of the variance.

### 3.1. Interactions

GDP, urban population, and population density, somewhat surprisingly, can be dropped without seriously affecting the overall model fit in Model 7. The stepwise model helps explain this, by showing that the interactions between these terms are nearly as important as the main effects. As previously mentioned, we should expect the dynamics of COVID-19 to be exponential or multiplicative. From this perspective, interactions match the dynamics we’re expecting. In isolation, each factor contributes only somewhat to the spread of COVID-19, but in combination, they are much more dangerous than the sum of their parts, and combinations of combinations are even more dangerous, etc.

As one example, population density would influence the spread of COVID-19 a lot more in a national population of 80-year-olds than it would in a national population of 9-year-olds. This has been observed on a smaller scale in that many superspreader events have occurred in elderly care homes, but rarely occur in elementary schools (Leclerc et al., 2020, Munro & Faust, 2020). As a result, it’s not surprising that an interaction term is a good way to model these dynamics, which might not appear in a main effect. The stepwise model can be interpreted as evidence in favor of the kind of exponential growth we should see from factors influencing a pandemic, which further supports the selection of these variables.

### 3.2. Proxies

One concern in modeling approaches is that the critical variables may be proxies for something else. There are two potential ways in which variables can be proxies in the situation presented here, and both appear unlikely.

The first possibility is that some or all of our variables — median age, obesity rate, island status, strict border closure policies — might be proxies for a single underlying variable. In this case, the natural concern is that they might be a proxy for something like wealth or development. After all, variables like age and obesity are moderately correlated with wealth. Our findings offer strong evidence that the critical variables are not functioning as proxies. First, we included GDP as a covariate in our models, so that if our critical variables were proxies for wealth and development, then GDP would compete with them in explaining variance. That’s not what we observe. Instead, the critical variables explain a large amount of additional variance (usually about 10%) above and beyond GDP. When all the critical variables are in a single model, GDP is no longer a significant predictor, and can be dropped without seriously reducing the fit of the model. In other words, they can account for the influence of GDP, but GDP can’t account for them.

Second, if our critical variables were all proxies for the same underlying variable (like wealth) they wouldn’t have statistical interactions with one another. Multiple measures or proxies of the same variable don’t have statistical interactions, because redundant measures don’t have multiplicative effects.

Now, if our variables are not all proxies for a single grand variable, they could still be proxies for variables individually; that is, they might accidentally be proxies for different variables that are more directly predictive of COVID-19 case spread. For example, median age could be a proxy for the rate of hypertension in a country, and obesity rate for the rate of physical activity. We included covariates such as GDP per capita PPP and measures of population density, but it’s possible that we missed a more relevant covariate.

Fortunately, there is also strong evidence that our variables are not proxies for individual concepts either. Interactions are relatively fragile, and three-way interactions especially so. If two variables have a statistical interaction, the interaction between close proxies for those variables will be reduced or even eliminated. This is especially true for three-way interactions, and in relatively small sample sizes, like the sample we have here. If these variables were proxies for something else, it’s unlikely that there would be three-way interactions including these variables; since we do observe these interactions, it strongly suggests the variables are not proxies for unknown confounding variables.

The fact that island status, border closure, median age, and obesity rate have multiple three-way interactions with each other, and with other expected predictors, is strong evidence that these variables are actual predictors of interest, rather than proxies for other variables.

### 3.3. Main Findings

Age has previously been found to be an important predictor of variance in COVID-19 rates between different countries (Iyanda et al., 2020; Gardiner et al., 2021), and unsurprisingly we replicated this finding with median age in our analysis. Somewhat surprising is that we found a slightly greater effect of age than was found in most other papers. This may be due to the fact that our data were collected later and factors that influence transmission should become more apparent over time, because we examined a larger set of countries than most previous analyses, or because the inclusion of other important predictors allowed median age to act as a better predictor itself.

Obesity rates, on the other hand, have not generally been seen as a major factor in explaining the differences in COVID-19 rates worldwide. Obesity has occasionally been found to be a significant predictor, but generally in limited circumstances, such as when examining the variation in a small subset of nations (Gardiner et al., 2021) or for a specific range of BMI values (Li et al., 2021). Our results strongly suggest that obesity has previously been overlooked because there is a critical interaction between obesity and island nation status that is easy to miss. Simply put, while there is a strong positive relationship between obesity rates and COVID-19 cases in mainland nations, the same relationship does not appear in island nations. In fact, this effect is exacerbated by the existence of a small number of island nations that are extreme outliers on both obesity and COVID-19 rates. Together this means that the relationship between country-level obesity rates and COVID-19 cases, when treated as a zero-order correlation or a main effect, appears to be close to zero, and sometimes even appears to be negative. It is only when including the appropriate statistical interactions that the full predictive power of obesity as a factor is revealed, and these interactions are easy to miss for researchers who do not anticipate them.

Something similar appears to be the case for border closure measures. While these measures make sense intuitively, the empirical evidence is relatively mixed. There has been only one large cross-country analysis examining the effectiveness of various non-pharmaceutical interventions (NPIs) on COVID-19 growth rates (Duhon et al., 2020). This study found that most NPIs appeared to have little to no impact on the growth of COVID-19, and while international movement restrictions were the only NPI that appeared to have a potential effect, this effect only approached significance. Like obesity, our results suggest that the full impact of border closure measures can only be understood by examining its interactions with other variables, especially its interaction with island status. Like obesity, border closure has a different impact on mainland and island nations. Island nations appear to benefit immensely from border closure measures, while mainland nations may not see any benefit at all.

Island nations that combine strong border closure with other aggressive COVID-19 policies seem able to reduce COVID-19 rates reliably and effectively (Leal Filho et al, 2020; Khana & Well, 2021). While this study did not evaluate the effectiveness of COVID-19 policy interventions other than border closure, it does appear possible that Island nations may be able to prevent the spread of COVID-19 with strong policies even if their population has a high median age and rate of obesity. Many island nations with a combined high median age and obesity rate had COVID-19 case rates below 1% in March of 2021, including Australia, New Zealand, Micronesia, Samoa, Marshall Islands, Fiji, Cuba, Trinidad and Tobago. Mainland nations with a similarly high median age and obesity rates did not do nearly so well. In fact, the only nations in the world which are universally accepted^2^ to have entirely avoided COVID-19 infections are the nations of Palau, Tuvalu, Kiribati, Nauru, and Tonga (Stephens, 2021), all island nations who kept strict border closure throughout the pandemic. Interestingly, during the 1918 influenza pandemic, the only nations to entirely avoid the pandemic were island nations with strong border closure policies, such as American Samoa (Patterson & Pyle, 1991). These extreme examples offer further support that the geographical isolation of island nations, in conjunction with strong border closure, creates an environment unique in its ability to protect against infectious disease pandemics such as COVID-19.

The importance of population age, obesity, and island status can help explain multiple worldwide patterns of COVID-19. For example, while COVID-19 was expected to be devastating in Sub-Saharan Africa (El-Sadr & Justman, 2020; Nkengasong, & Mankoula 2020), exponential growth never occurred in these nations, even after infections were identified. Other than South Africa, every Sub-Saharan nation had a cumulative COVID-19 rate well below 1% by March of 2021. South Africa is a notable exception with much higher COVID-19 rates (> 2.5% of the population), but it is also notable that South Africa has the highest median age and obesity rates in mainland Sub-Saharan Africa.

The low rates of COVID-19 in East and Southeast Asia have typically been credited to aggressive COVID-19 policies, which certainly did reduce COVID-19 transmission. However, the relatively low obesity rates in these countries may further explain the dramatic differences in COVID-19 rates when compared with European nations. Countries which have been credited with an excellent COVID-19 response include Vietnam, Thailand, Finland and Norway (Djalante et al., 2020; Christensen & Lægreid, 2020; Kabiraj, & Lestan, 2020), yet COVID-19 rates in Finland and Norway are at least 30 fold higher than Thailand and over 100 fold higher than Vietnam. It is unlikely that these large variations in COVID-19 rates between these nations is fully explained by policy differences, given cross-country analysis have failed to consistently identify policies with such a dramatic impact (Bendavid et al., 2021).

Policy interventions and other non-pharmaceutical interventions (NPIs) appear to have an inconsistent impact on the spread of COVID-19 within and between nations (Duhon et al., 2021, Chin et al., 2021, Savaris et al., 2021, Bendavid et al., 2021). This an ongoing mystery, given that these interventions are based on firmly established principles of infectious disease control (Collett et al., 2020). Given that age, obesity rate, island status, and their interactions explain much of the variation in COVID-19 rates between nations, inconsistent effects of NPIs should be expected if these variables are not accounted for in cross-country comparisons. Border closure measures are a strong example of this, as they appear to have a strong effect in island nations, and appear to have little or no effect in mainland nations, but this is not revealed unless one examines the relevant interactions. Future modeling studies must account for population age, obesity, island status, and border closure as well as any interactions, to establish more consistent results and correctly identify the policies which are most effective.

Many academic and press articles have attempted to explain why countries have experienced a high or low rate of COVID-19, frequently identifying interventions the country had employed or failed to employ (Jones 2020, Douglas, 2020). While some interventions can reduce COVID-19 rates, interventions do not appear to be the predominant cause of the variation in COVID-19 rate observed worldwide. Our findings strongly suggest that median age, obesity rate, island status, and border closure explain the majority of the variance of COVID-19 rates worldwide, and models based on these variables perform better than previous attempts.

The findings of this study may offer guidance for future policy decisions, centered on each nation’s median age, obesity rate, and status as an island. Border closure measures, for example, seem particularly important for island nations, and largely unimportant for mainland nations. Obesity rates, on the other hand, are a good predictor of the severity of an outbreak in mainland nations, but not in island nations. Many policies tailored to control COVID-19 rates, such as business closures and stay at home orders, threaten to cause economic and psychological harm, especially among the poorest nations (Piper, 2020). When attempting to balance the benefits and harms of any policy, nations with low median age and obesity rates, should consider less harmful policies that can be put in place without a devastating result on their COVID-19 rates. It would be prudent for any nation with low median age and obesity rates to minimize the most harmful COVID-19 policies, and to continue to bar entry from nations with high COVID-19 rates if it is an island.

These dynamics are likely to change over time, with the introduction of vaccines, as the virus mutates, and as different strains of the virus undergo selective pressure. The role of median age and obesity rate are likely to decrease given worldwide disparities in COVID-19 vaccine distribution to wealthier nations (Rouw et al., 2021). As the virus mutates, any variant with a mutation that causes higher susceptibility or increased transmission in younger populations or populations with lower rates of obesity will have a notable evolutionary advantage, and likely spread. A variant of this nature could cause a catastrophic resurgence of COVID-19 in much of the developing world, countries which have been protected by their relatively low median age and low obesity rates. In fact this may already be occurring, with the Delta variant first observed in India being associated with unprecedented rises in daily cases of COVID-19 in Asia and Africa (Mendez, 2021). This possibility raises the importance of variant surveillance, specifically examining enhanced susceptibility or transmissibility based on age and BMI. On the other hand, an examination of historical evidence suggests that the role of island status and border closure, and especially their combined effect as found in maritime quarantines, is likely to be apparent in future pandemics of any kind.

## Data Availability

All data is available at the listed OSF repository page

https://osf.io/prm6y/

## Acknowledgements

The authors would like to thank Adam Mastroianni, Siavash Sarlati, Brian Piper and Grace Rosen for helpful discussion and comments.

## Footnotes

1 The Pacific Island nations of Palau, Tuvalu, Kiribati, Nauru, and Tonga, are universally accepted to have experienced no COVID-19 infections to date. The nations of North Korea and Turkmenistan report having had no cases of COVID-19, however given the authoritarian regimes of these countries there is reason to suspect the authenticity of these claims (Stephen, 2021).

2 North Korea and Turkmenistan also have no reported cases; however, given the authoritarian governments of these two nations, it is generally believed that COVID-19 cases have been identified but not publicly reported (Stephen, 2021).

